# Rare GATA6 variants associated with risk of congenital heart disease phenotypes in 200,000 UK Biobank exomes

**DOI:** 10.1101/2021.05.04.21256616

**Authors:** Simon G. Williams, Dominic Byrne, Bernard D. Keavney

## Abstract

Several genes have been associated with congenital heart disease (CHD) risk in previous GWAS and sequencing studies, but studies involving larger numbers of case samples remain needed to facilitate further understanding of what remains a complex and largely uncharacterised genetic etiology. Here we use whole exome sequencing data from 200,000 samples in the UK Biobank to assess ultra-rare and potentially pathogenic variation associated with increased risk of CHD. Our findings indicate that rare variants in GATA6, presumably with a lesser effect on gene function than those causing severe CHD phenotypes, or buffered by other genetic and environmental effects during development, are also associated with minor CHD conditions, specifically bicuspid aortic valve, the most common CHD condition.

## Research Letter

Anomalies arising in early embryonic development can result in a range of congenital heart disease (CHD) phenotypes, from complex conditions with multiple defects, such as tetralogy of Fallot (TOF), to comparatively mild phenotypes that may go undiagnosed until later life, such as isolated bicuspid aortic valve (BAV) [1]. Several genes have been associated with CHD risk in previous GWAS and sequencing studies, but studies involving larger numbers of case samples remain needed to facilitate further understanding of what remains a complex and largely uncharacterised genetic etiology.

Here we use whole exome sequencing (WES) data from the first 200,000 samples in UK Biobank (UKB) to assess rare and potentially pathogenic variation associated with increased risk of CHD. Using hospital episode statistics (HES) and primary care diagnostic codes in a classification scheme we previously reported [2], we identified 1354 individuals with CHD phenotypes and classified 179310 individuals as non-CHD controls. The most common CHD-related codes identified were aortic stenosis (n=395), aortic insufficiency (n=278) and evidence of aortic valve replacement (n=270). We inferred that clinically manifest aortic valve disease (AVD) before the age of 65 was highly likely to be due to BAV, and classified cases accordingly. UKB participants with AVD manifesting only after age 65 were assigned unknown status. As previously observed, more severe CHD conditions such as atrial septal defect (n=76), ventricular septal defect (n=69) and TOF (n=13) occur infrequently in UKB participants, demonstrating the known ‘healthy cohort bias’ in UKB where more severe CHD phenotypes are under-represented. The UKB cohort therefore represents the largest case/control analysis of predominantly mild CHD phenotypes with WES to date.

We conducted a variant analysis to identify increased burden of ultra-rare, potentially pathogenic variants at the gene level in CHD cases. Variants were first filtered (genotype quality (GQ) >=20; read depth (DP) >=10 for indels and DP>=7 for SNPs) before being annotated with Variant Effect Predictor (VEP) v98. To focus on potentially pathogenic variants, we retained only variants present in <1% of the total UKB samples, with HIGH/MODERATE impact, absent in gnomAD and CADD score >=20. Additionally, to assess potential differences in more common variation between case and control groups, synonymous variants with gnomAD AF>0.01 were extracted. Qualifying variants (QV) for each analysis were collapsed to the gene level for burden analysis.

Following removal of related individuals and ensuring ancestral similarity between groups using principal component analysis, CHD case and control cohorts were compared. For common variants, the QQplot of variants (figure 1A) indicates no difference between case and control groups. Conversely, the QQplot of ultra-rare pathogenic variation (figure 1B) shows inflation caused by a small number of genes with significantly increased prevalence in the CHD case group. A burden analysis of these rare pathogenic QV highlights GATA6 as the gene with the most significant association with CHD (OR=7.66 [95% CI 3.6-14.47]; p=1.60E-06), with 9 unique QV present in 10 CHD samples, and the only significant candidate following Bonferroni correction (p=0.03).

**Figure 1.**
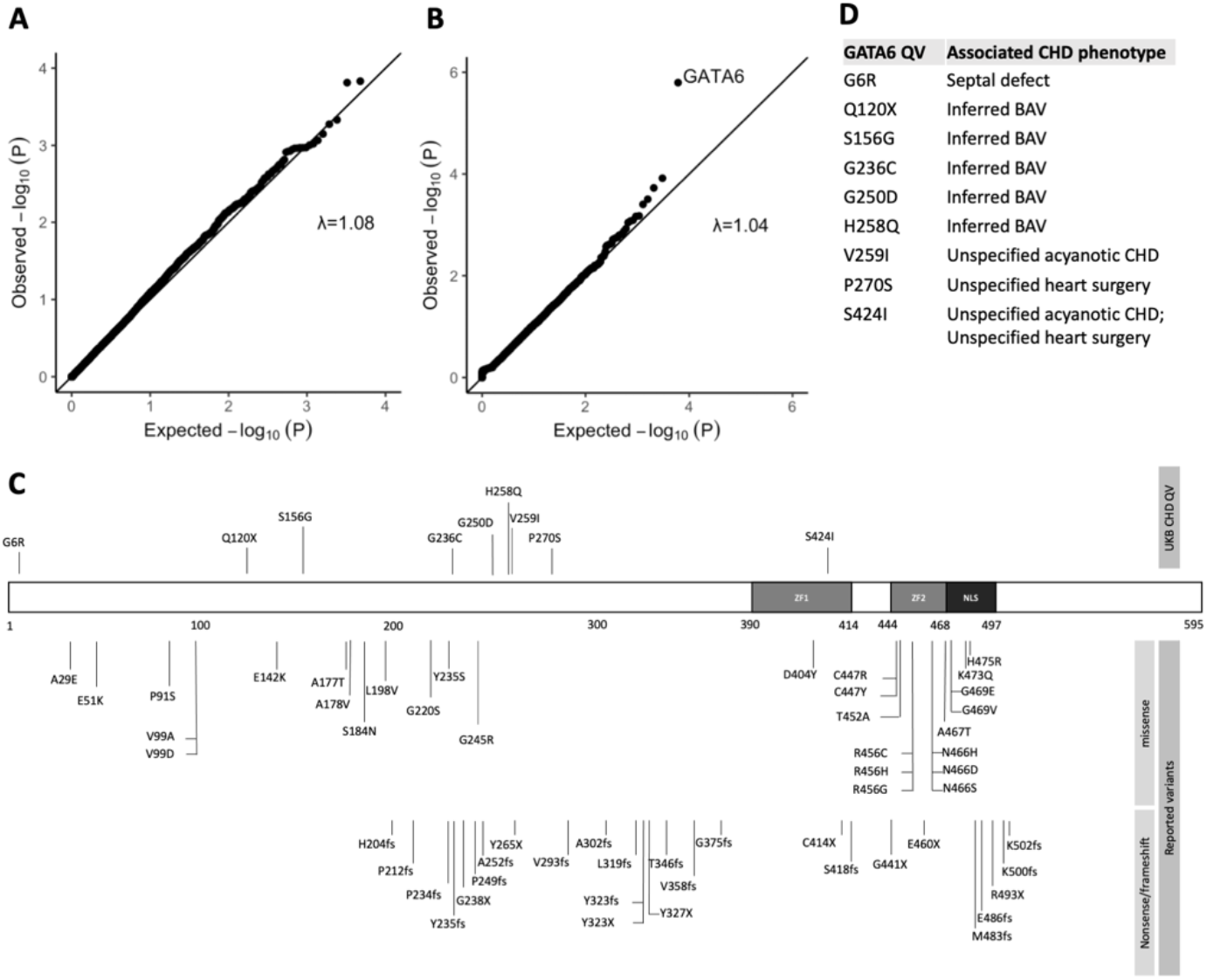
**A**. QQ plot of common (gnomAD AF>0.01) synonymous variants between CHD cohort and controls in autosomes. **B**. QQ plot of rare (not present in gnomAD) and potentially pathogenic (HIGH/MODERATE impact and CADD>=20) variants between CHD cohort and controls. **C**. Schematic of GATA6 protein with two DNA-binding zinc-finger domains (ZF1, ZF2) and nuclear localisation sequence (NLS). The location of previously reported variants – both missense and nonsense/frameshift – are shown along with the UKB CHD QV. **D**. The CHD phenotypes of the individuals with GATA6 QV.

GATA6 is a member of a family of zinc-finger transcription factors that play an important role during heart development by regulating cellular differentiation. GATA6 has previously been associated with a range of CHD phenotypes, the first association described being with persistent truncus arteriosus, though association with BAV has not previously been shown in population genomic data. Mice haploinsufficient for GATA6 have been shown to have BAV through a proposed mechanism of dysregulated extracellular matrix regulation, and human BAV tissues removed at surgery have lower levels of GATA6 expression than do tricuspid aortic valves [3]. GATA6 has a similar expression pattern to GATA family member GATA4, a well-established CHD-associated gene; a previous GWAS showed association between protein-altering and regulatory common variants near GATA4 and BAV [4]. A recent paper [5] reviewed the literature on the genotypic and phenotypic spectrum of GATA6 in humans, with focus on more severe conditions. Eighty percent of mutation carriers reported structural cardiac phenotypes, predominantly atrial or ventricular defects. Additionally, the association of GATA6 with a range of pancreatic anomalies was confirmed. Previously reported CHD-associated missense variants cluster in the DNA-binding domains where they likely severely disrupt function. Mapping the UKB CHD QVs to the GATA6 protein shows no clustering in these domains (figure 1C), indicating that some GATA6 function may be retained, potentially resulting in the milder CHD phenotypes observed (figure 1D). Additionally, the case samples with GATA6 QVs have no previous history of pancreatic anomalies.

Our findings indicate that rare variants in GATA6, presumably with a lesser effect on gene function than those causing severe CHD phenotypes or buffered by other genetic and environmental effects during development, are also associated with minor CHD conditions, specifically BAV. BAV affects around 1% of males and 0.33% of females and is therefore the commonest CHD condition. The absence of other gene candidates in this study confirms the wide heterogeneity of CHD phenotypes where rare variants in single genes can only account for a small proportion of cases.

## Data Availability

Data used in the manuscript are available from UK Biobank

## Acknowledgements

This research has been conducted using the UK Biobank Resource under project 19056 and was supported by the British Heart Foundation Programme Grant RG/15/12/31616.

## References

1. Liu, Y., S. Chen, L. Zuhlke, et al., Global birth prevalence of congenital heart defects 1970-2017: updated systematic review and meta-analysis of 260 studies. Int J Epidemiol, 2019. 48(2): p. 455–463.

2. Williams, S.G., A. Nakev, H. Guo, et al., Association of congenital cardiovascular malformation and neuropsychiatric phenotypes with 15q11.2 (BP1-BP2) deletion in the UK Biobank. Eur J Hum Genet, 2020. 28(9): p. 1265–1273.

3. Gharibeh, L., H. Komati, Y. Bosse, et al., GATA6 Regulates Aortic Valve Remodeling, and Its Haploinsufficiency Leads to Right-Left Type Bicuspid Aortic Valve. Circulation, 2018. 138(10): p. 1025–1038.

4. Yang, B., W. Zhou, J. Jiao, et al., Protein-altering and regulatory genetic variants near GATA4 implicated in bicuspid aortic valve. Nat Commun, 2017. 8: p. 15481.

5. Skoric-Milosavljevic, D., F.V.Y. Tjong, J. Barc, et al., GATA6 mutations: Characterization of two novel patients and a comprehensive overview of the GATA6 genotypic and phenotypic spectrum. Am J Med Genet A, 2019. 179(9): p. 1836–1845.

